# Oral injury in Kendo players: a cross-sectional survey

**DOI:** 10.1101/2022.03.18.22272564

**Authors:** Koshiro Watanabe, Gen Tanabe, Hiroshi Churei, Takahiro Wada, Yuka I Sumita, Kunihide Koda, Kichio Uehara, Toshiaki Ueno

## Abstract

This study evaluated the risk of oral injury in Kendo. We hypothesized that Kendo players in Japan may experience oral injury due to the use of face protector for kendo (*men*) with inaccurate measurements or wearing it incorrectly. The survey included 400 kendo players (male, 276; female, 174) and covered four areas: the relationship between the characteristics, percentage of oral injury, temporomandibular disorder (TMD), and use of *men*. Based on the who experience of oral injury, participants were classified into ‘trauma’ and ‘non-trauma’ groups. Those who suffered oral injury were 179 (44.8 %; males, n=118; females, n=61). In the past month, 32 (8.2%) Kendo players reported that their matches/training were affected by dental/oral problems. For TMD, 50 participants had a total score >8.5 from four screening questions. For 289 (72.3%) participants, the *men* they are currently wearing fit well. Of those that reported that their *men* do not fit properly, 48 (12.0%) felt that their *men* were too large, and 14 (3.5%) felt that their *men* were too small. Years of experience and clenching of one’s jaws during an offensive Kendo movement significantly contributed to oral injury. Clenching one’s jaws during a defensive Kendo move, current *men* fit, sleep bruxism, and morning symptoms from sleep bruxism significantly contributed to TMD. *Men* with an accurate sizing allows proper field of vision, eliminating head shifting inside *men* during strikes and movements. This suggests that vision can be secured by adjusting the ties on *men*, chin movement, and the way one looks through the *men*. Since Kendo equipment is originally a Japanese craft, it has high artistry. Thus, properly worn Kendo equipment brings higher functionality and esthetic appeal.

## Introduction

Kendo is a modern Japanese *budō* or martial way: *Budō* is a philosophy and a way of life that utilizes martial arts as a means of self-improvement. Since the 19th century, various styles in swordsmanship have been practiced all over the nation, and Kendo was established under the commitment of the Greater Japan Martial Virtue Society with the development of a specific rule in accordance with modernization in Japan [1]. Recently, it was estimated that approximately 2.5 million people in over 59 countries practice Kendo [2].

Movement in Kendo is similar to the western practice of fencing; however, kendo imbibes the unique essence of Japanese martial arts to train one’s mental and physical strength, based on its monistic philosophy [3,4]. Kendo players are referred as “*kendo-ka*” or “*kenshi*”; they use bamboo swords called *“shinai*” and wear protective gear called “*kendo-gu*”. There are four types of “*kendo-gu*”: the piece of kendo equipment which covers the head, face, throat, and shoulders (*men*), the glove which cover the hands and forearms (*kote*), the trunk protector which covers the chest and stomach areas (*do*), the hip protector which is worn around the waist, and which covers and protects the lower abdominal area and the thighs (*tare*). Modern Kendo techniques consist of both thrusts and strikes, targeting areas: the head (*men-bu*), the right forearm (*kote-bu*), torso or both sides of the body (*do-bu*), and throat (*tsuki-bu*) (Fig 1)

**Fig 1.**
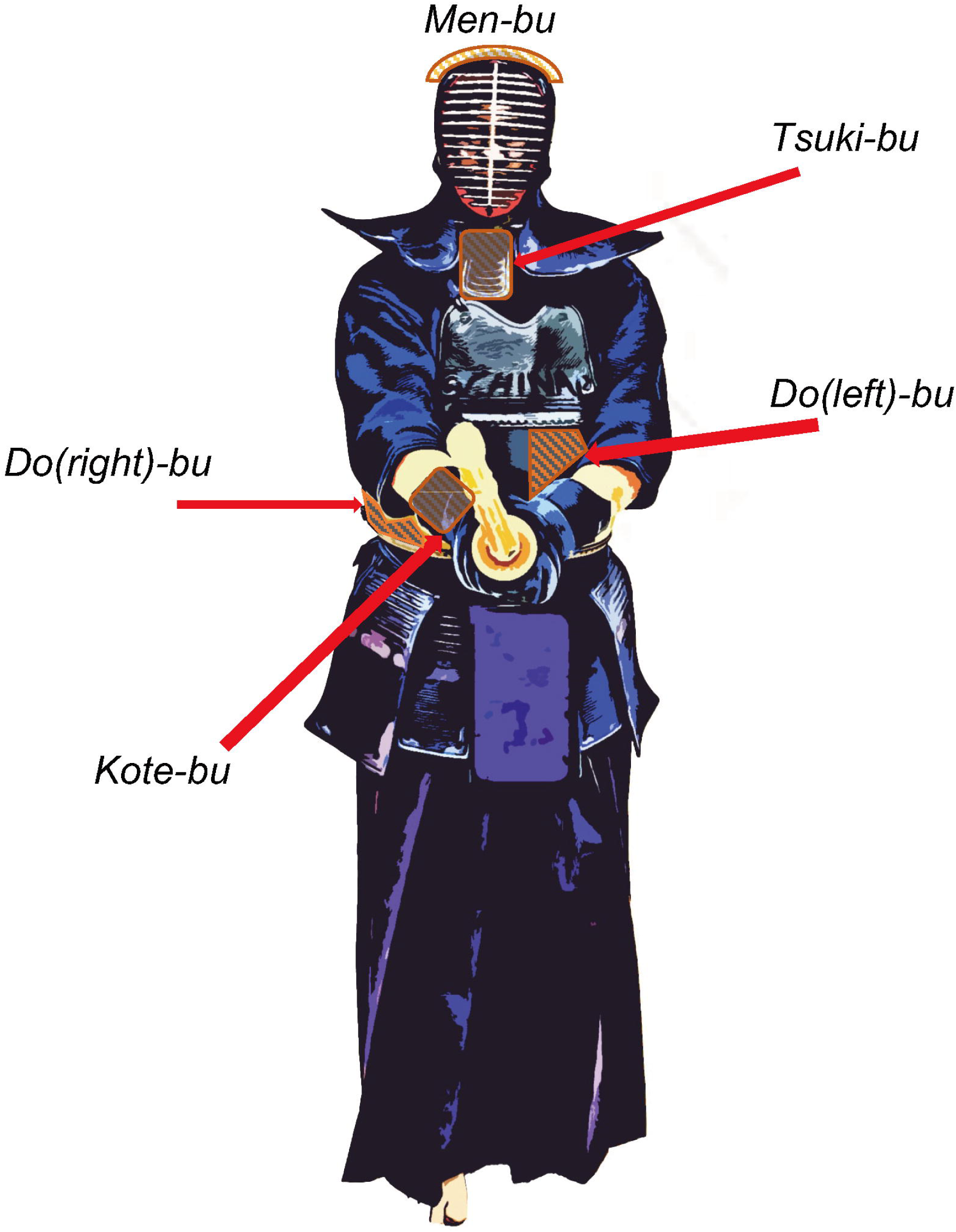
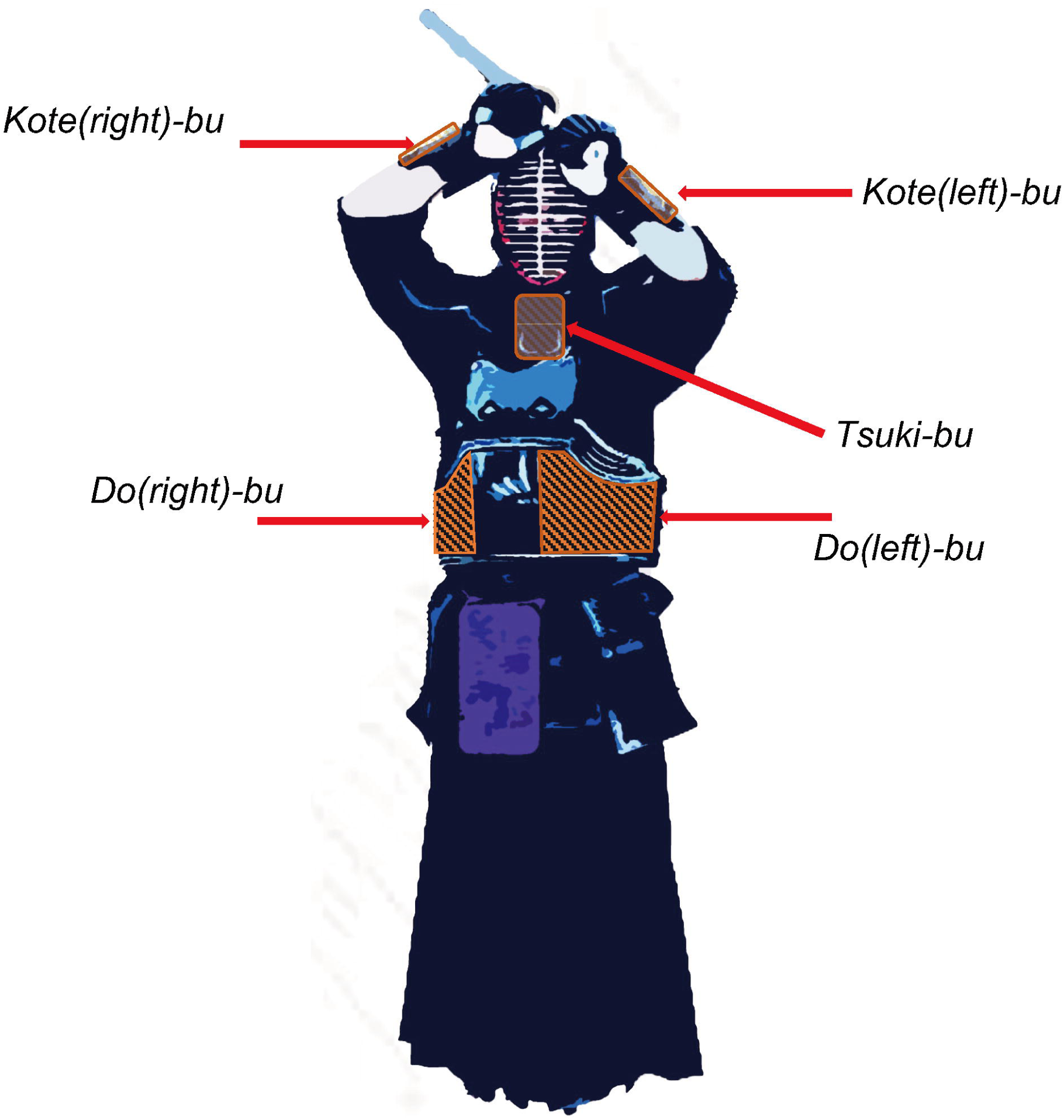
Targeting areas: General term for the designated striking zones of *men-bu, kote-bu, do-bu*, and *tsuki-bu*. When these targets are struck (or thrust) according to the requires criteria, it will be considered a valid point.

With increasing number of Kendo players, there has been an emphasis on the functionality of kendo equipment. Modifications to *shinai* and *kendo-gu* have led to a threat to competitors’ safety. In the regulations [5], there is a trend of lightening the armors to increase agility; for example, making *men* thinner by using less protective padding material inside and shaving out the inside of *the shinai*. These changes bring forth the question of safety. To tackle this rising concern, safety regulations require further study of injuries related to the modification of the equipment to establish regulations for its size and materials.

In 2019, the All-Japan Kendo Federation made a committee statement on amendments to the Match Referee Rules for Kendo Equipment for further safety [5,6]. This guideline sets rules to regulate the thickness of the tip of *the shinai* and the length of *Kendo-gi’s* sleeve, uniform, and a *kendo-gu* with a guaranteed cushion to decrease the impact while performing Kendo. However, this regulation does not consider teenage Kendo players’ hearing issues and injuries caused by strikes [7,8]. It was suggested that these issues may be due to inaccurate measurements of the gear or a lack of instruction leading to incorrect wearing of the gear, making it incapable to fully absorb the impact from the strikes [9].

These issues in wearing kendo gear and associated problems have been explored. Kato et al. demonstrated that many *Kendo-ka* do not wear matching *men*, especially children and performers who have studied outside of Japan, and also studied the relationship between the route of purchase and the eye position of the *men* [10]. No study has detailed the relationship between the use of Kendo equipment and oral injuries.

The purpose of this study was to investigate the prevalence of oral injury in Kendo players; further, the risk of dental injury based on the characteristics of Kendo and its equipment were assessed.

## Materials and methods

### Participants and data collection

Data were collected from 400 Kendo players with 100% response rate. The study sample consisted of male (n=276) and female (n=124) Kendo players in Japan, who were junior high school (n=101), high school (n=157), and university (n=142) students. Sex, age, height, weight, years of experience, number of practice days per week, and time spent wearing the protective *men* per day are shown in Table 1. The majority of them, 366 (91.5%), had practiced Kendo since elementary school.

**Table 1.** Characteristics of the participants.

|  | School category |  |  |
| --- | --- | --- | --- |
|  | Junior high<br>school students<br>(N = 101) | High school<br>students<br>(N = 157) | University<br>students<br>(N = 142) |
| Male / Female | 67 / 34 | 105 / 52 | 104 / 38 |
| Height (cm) | 158.1 ± 8.0 | 165.7 ± 7.1 | 168.7 ± 7.6 |
| Weight (kg) | 51.1 ± 10.3 | 63.0 ± 9.9 | 65.4 ± 9.1 |
| Years of experience (year) | 6.3 ± 2.9 | 9.4 ± 2.4 | 12.2 ± 2.5 |
| Practice days per week (day) | 5.9 ± 1.1 | 6.7 ± 0.6 | 5.8 ± 0.7 |
| *Men* (*men*) wearing time per day<br>(min) | 106.3 ± 49.4 | 135.3 ± 42.9 | 100.8 ± 23.2 |
| mean ± standard deviation |  |  |  |

This cross-sectional study was performed using an anonymous questionnaire through Google Forms (Google LLC, CA, USA) with the approval of the ethics committees of Fukuoka University (Fu-18-10-03) and Tokyo Medical and Dental University (D2018-069), Japan. The purpose and contents of the study questionnaire were explained before the study commenced, and informed consent or representative approval was obtained from the participants and their parents. Questionnaires were then sent to all participants, and completed questionnaires were collected using a spreadsheet on a Google server.

### Questionnaire

The questionnaire focused on four areas: 1) attributes, 2) prevalence of oral injury, 3) relation to temporomandibular disorder (TMD) screening, and 4) usage of *men* equipment. To better understand the status of oral injury in Kendo, questions about the site of the injury, the situation at the time of the injury, use of mouthguards, and jaw clenching when playing Kendo were added to the questions on rate of injury.

Questions about clenching were scored a 5-point numeric rating scale (1–5). Scores 1 and 2 were grouped into “no clenching”, and scores 3–5 was grouped into “clenching”. Kendo player with oral trauma experiences were assigned to a “trauma” group, whereas those without oral trauma experiences were assigned to a “non-trauma” group.

Questions related to screening for TMD were based on a study by Sugisaki et al. [11]. The participants rated the four screening items using a 5-point numeric rating scale (1–5): one item about temporomandibular joint (TMJ) sounds; three items assessing psychosocial factors, including stress, anxiety, and depressed mood; and six items related to habitual behavior, including teeth-contact habits and morning symptoms that presumably resulted from sleep bruxism. The sum of the scores of the four screening items was used for risk screening of TMD with cut-off value of 8.5: participants with a score ≥9.0 were assigned to the “high risk of TMD” group, whereas those with a score <≤8.0 were assigned to the “low risk of TMD” group.

The questions about *men* included the current and past fit of respondents’ *men*, the position of one’s *men* in relation to their face and eyes (Fig 2), the tension of the knot securing the *men* on the face and methods of dealing with ill-fitting *men*.

**Fig 2.**
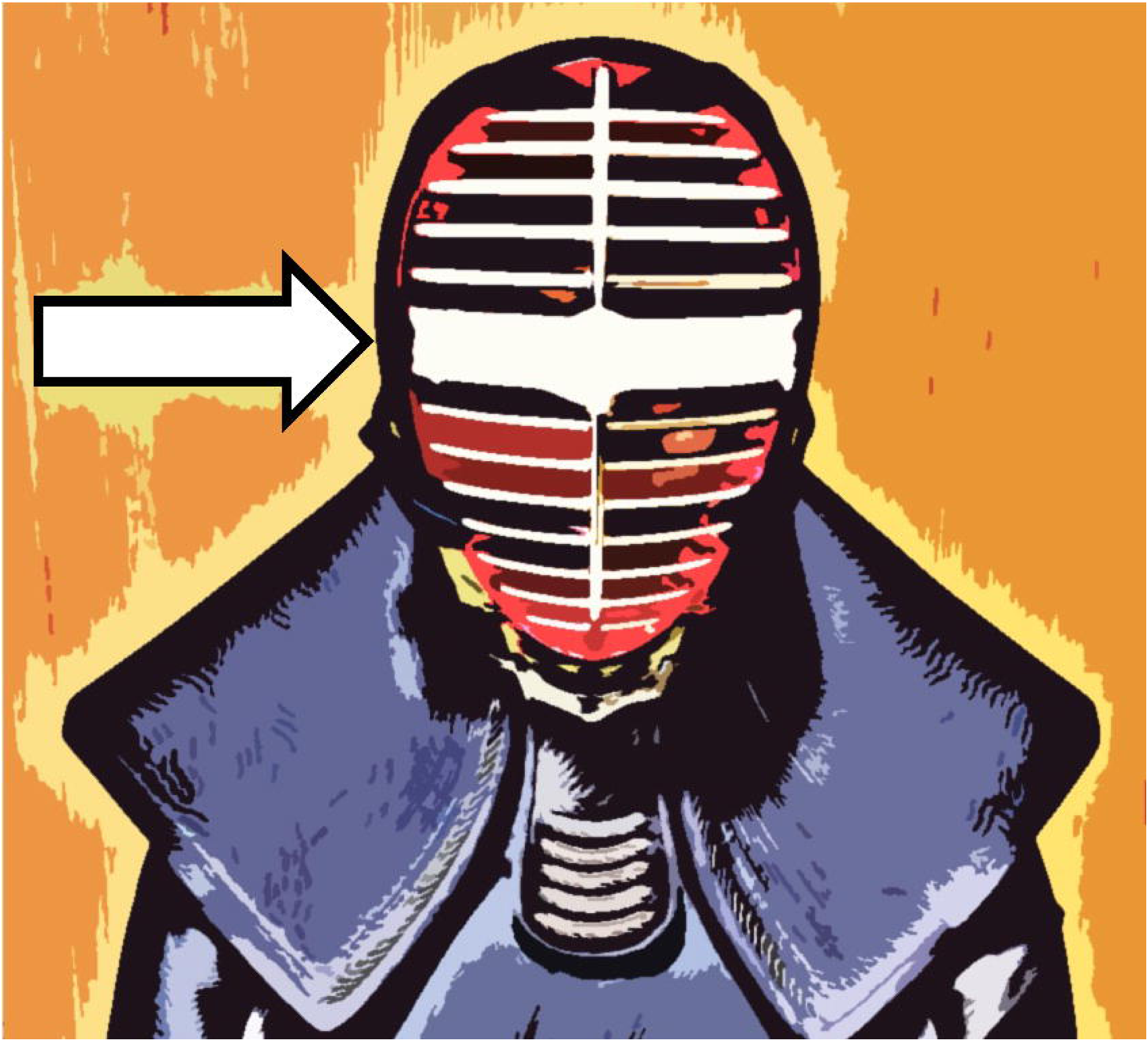
*“Monomi”*: a specific part of the protective*men* where the distance between the protective metal bars (“men-gane”) at the front of the men has been increased to allow the wearer better visibility. The white band in the picture shows the space between the sixth and seventh men-yokogane, the horizontal mental bars on the men. This space is a little wider than the others. It is important to puton the men so that the opponent can be viewed through this space.

### Statistical analysis

All recorded values were tabulated using JMP^®^ 14 (SAS Institute Inc., Cary, NC, USA) for statistical analyses. Data is presented as mean ± standard deviation.

The statistical analysis compared the traumatic groups and non-traumatic groups or high risk of TMD group and low risk of TMD group using chi-squared tests and Wilcoxon signed rank tests about each recorded value. Statistical significance was considered at a P value of less than 0.05. Subsequently, variable selection was performed using a stepwise method with accounting for multicollinearity, and these were used as independent variables in logistic regression analysis to obtain odds ratios (ORs) and 95% Cis as measures of association.

## Results

### Prevalence of Oral Injury

In Kendo player surveyed, the total number of students who suffered oral injury was 179 (44.8 %), with the number of males being 118 and females being 61. In the past month, 32 (8.2%) Kendo player that their matches or training had been affected by dental or oral problems in the past month. The most frequent site of intraoral injury was mucosal soft tissue (76.5%), followed by the TMJ (17.8%), with only 1.9% of injuries on hard tissues such as teeth or bone. Some Kendo players had severe dental trauma: six reported crown fracture, and two reported tooth luxations. Of the 179 who reported oral trauma, 58 (32.4%) occurred during a Kendo attack and 92 (51.3%) experienced a Kendo defense. Oral trauma also occurred in *kakari-geiko*, a specific high-intensity training exercise in Kendo, with 96 (53.6%) occurring in the aggressor who delivered a body-to-body blow (*tai-atari)*, and 101 (56.4%) in the receiver. Only three players (0.8%) reported using mouthguards on a regular basis.

### TMD

The distribution of the TMD screening test scores is shown in Fig 3. The high risk of TMD group, with scores ≥9.0 on the four-question screening test, had 50 participants (12.5%; males, 30 [10.9%] and females, 20 [16.1%]). Further, 114 participants reported temporomandibular joint sounds (28.5%), and 24 (6%) reported it every time they opened or closed their mouths.

**Fig 3.**
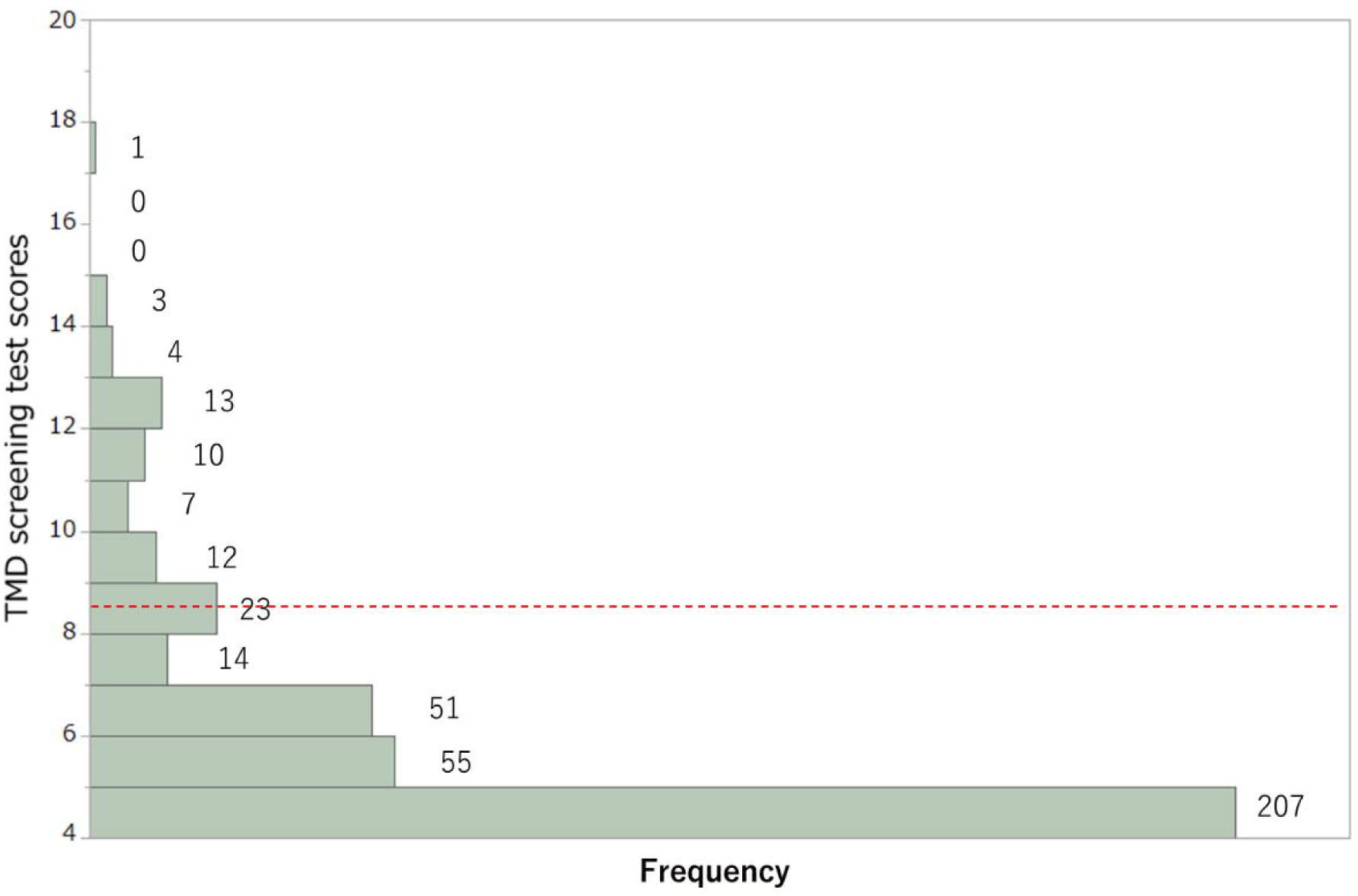
Distribution of the TMD screening test scores. Participants with scores ≥9.0 were assigned to the “high risk of TMD” group, whereas those with scores ≤8.0 were assigned to the “low risk of TMD” group. TMD, temporomandibular disorder

TMD, temporomandibular disorder

### Equipment /*men*

In total, 289 (72.3%) participants reported that the *men* they are currently using fits their face properly, while 111 (27.7%) felt that it did not fit. Of those that reported their *men* do not fit properly, 48 (12.0%) felt their *men* too large, and 14 (3.5%) felt their *men* too small. Some Kendo players also complained of reasons for ill-fit, unrelated to size. To counteract poorly fitting *men*, participants reported using a towel, an additional padding insert for the top of the face mask, a customized cloth chin sweat absorber placed inside the face mask or adjusting the tightness of the thin rope (*men-himo*) holding the face mask. The relationship between these measures to counteract oral injury and suspected TMJ disorders is shown in Tables 2 and 3.

**Table 2.** Relationship of each countermeasure to the trauma groups and non-trauma groups.

|  | Groups |  |  |
| --- | --- | --- | --- |
|  | Trauma | Non-trauma | p-value |
|  | (N=179) | (N=221) |  |
| Towel, a padding insert for the top of the face mask<br>(use/ no use) | 26/153 | 30/191 | 0.23 |
| Chin sweat absorber | 11/168 | 14/207 | 0.93 |
| Adjusting the tightness of *men-himo* | 23/156 | 31/190 | 0.73 |

**Table 3.** Relationship of each countermeasure to suspected TMJ disorder.

|  | Groups |  |  |
| --- | --- | --- | --- |
|  | Low risk of | High risk of | p-value |
|  | TMD<br>(N=350) | TMD<br>(N=50) |  |
| Towel, a padding insert for the top of the face mask<br>(use/ no use) | 48/302 | 8/42 | 0.66 |
| Chin sweat absorber | 22/328 | 3/47 | 0.54 |
| Adjustment to the tightness of *men-himo* | 47/303 | 7/43 | 0.91 |

TMJ, temporomandibular joint

Among the participants, 265 (66.3%) responded that the position of their *men* in relation to their faces and eyes was correct, 135 were aware of misalignment, 82 (20.5%) had their eyes positioned too low in the *men*, 53 (13.3%) had their eyes positioned too high, while 238 (60.0%) players reported tying an exceptionally tight knot to secure their *men*. Further, 366 (91.5%) players started wearing *men* for the first time in elementary school and the average number of times they changed the *men* was 3.6 ± 1.7 times, with the time of replacing *men* likely to be at school admission rather than due to ill-fit. In total, 265 (66.3%) Kendo players answered that the *men* they wore in the past did not fit their face size. Of these, 231 (57.8%) felt that their *men* were too large, 136 (34.0%) felt that their *men* were too small, and 102 had issues with both large and small *men* in the past. To counteract poorly fitting *men* in the past, the measures used were similar to the aforementioned methods, such as using a towel, a customized cloth chin sweat absorber, and adjusting the tightness of the thin rope (*men-himo*),and new *men* were bought by 40 (17.3%) of those who used larger *men* and 46 (33.8%) of those who used smaller *men*.

### Factors associated with oral injury and TMD

Tables 4 and 5 show the results of chi-squared tests and Wilcoxon signed-rank tests for each recorded value to compare between the trauma and non-trauma groups and the high risk and low risk of TMD groups, respectively.

**Table 4.**
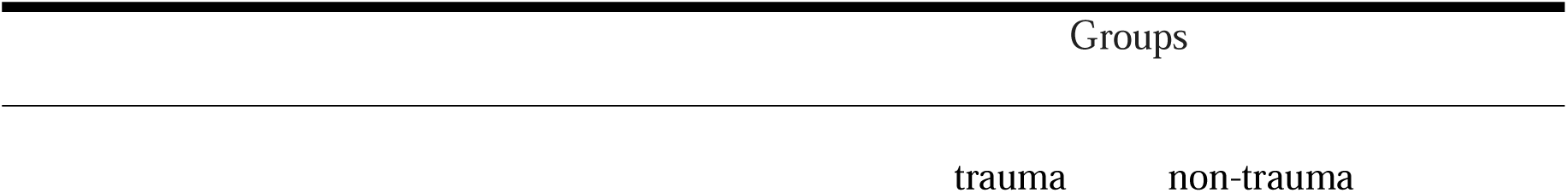
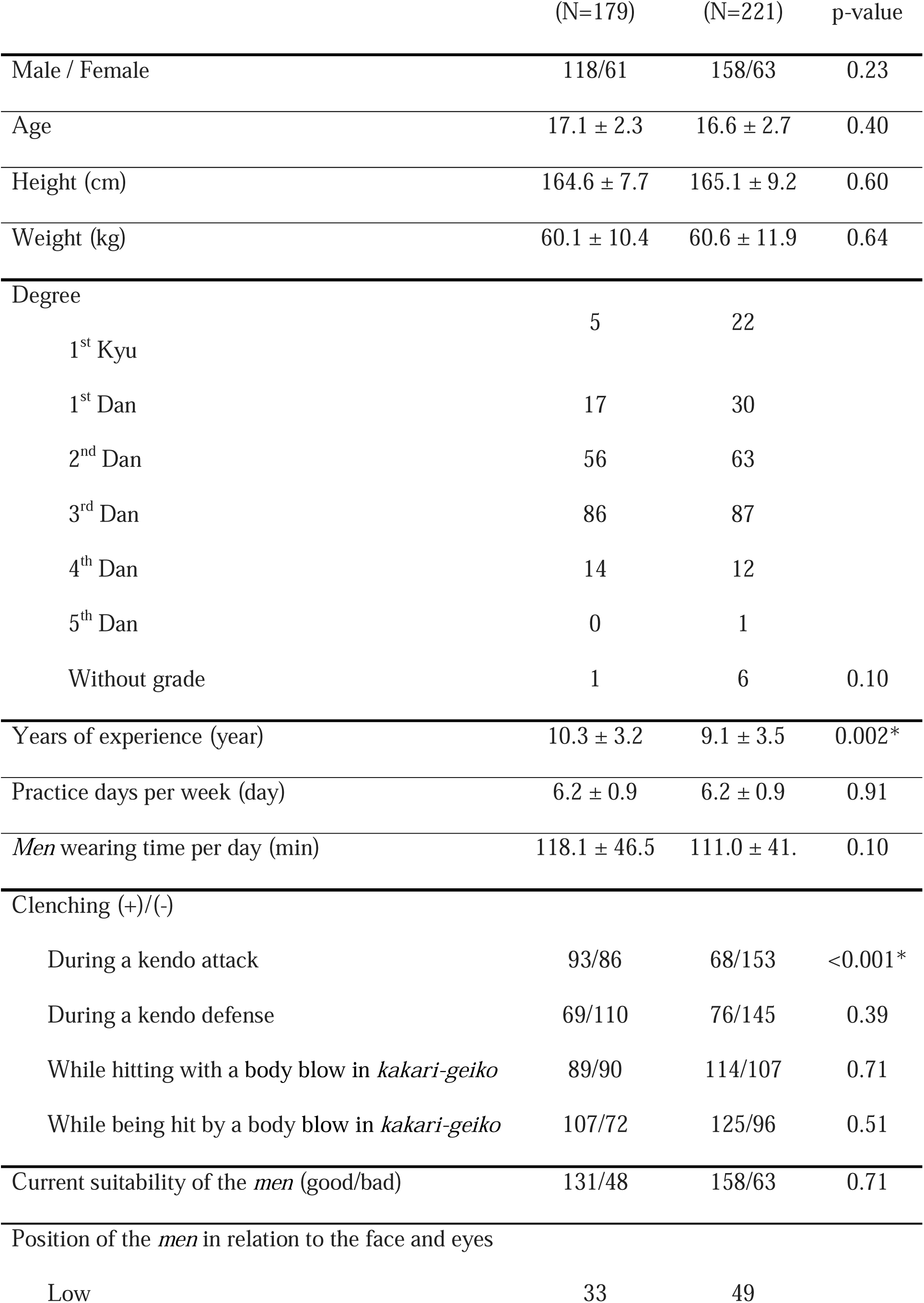
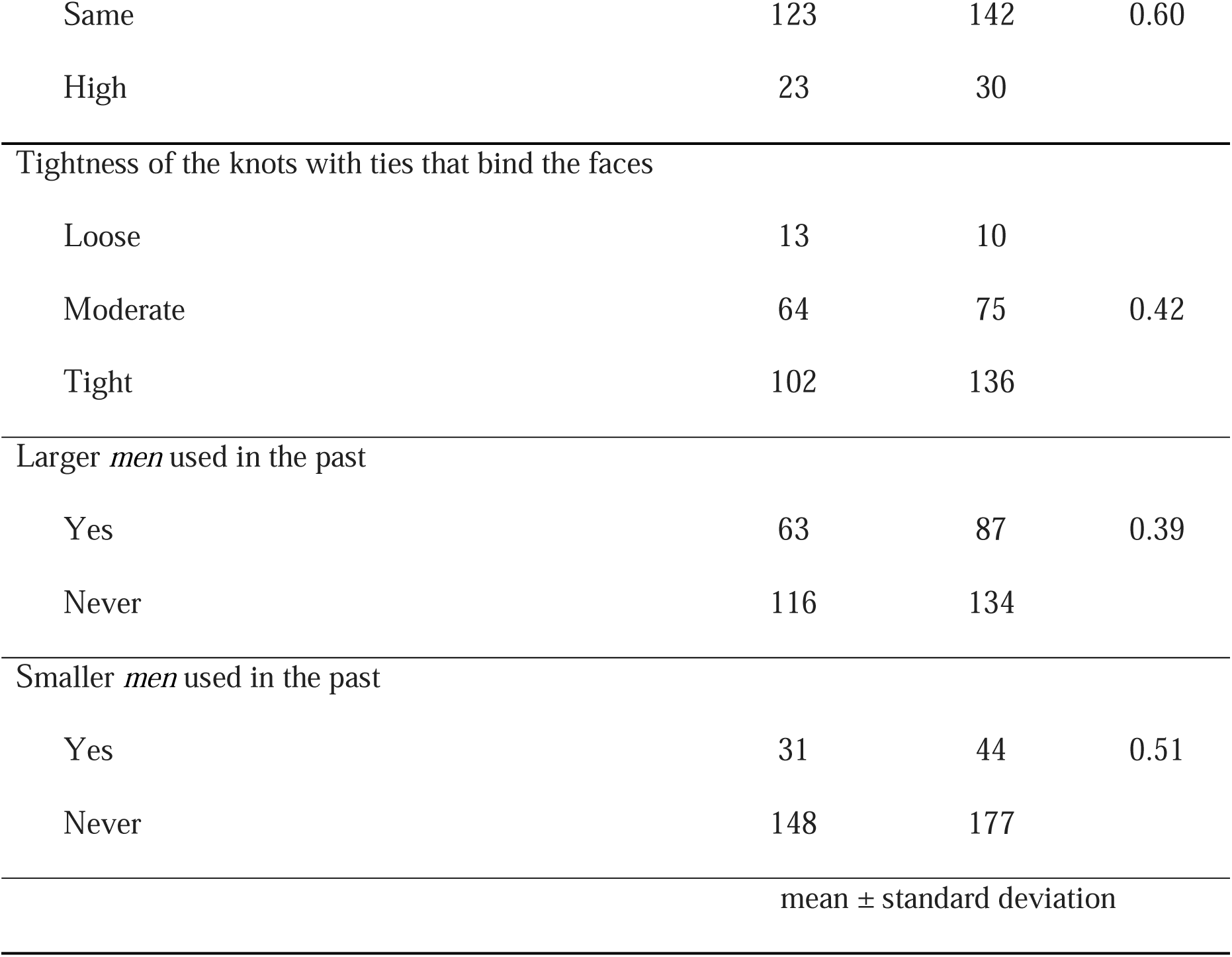
Characteristics of the total participants according to trauma and non-trauma groups.

**Table 5.**
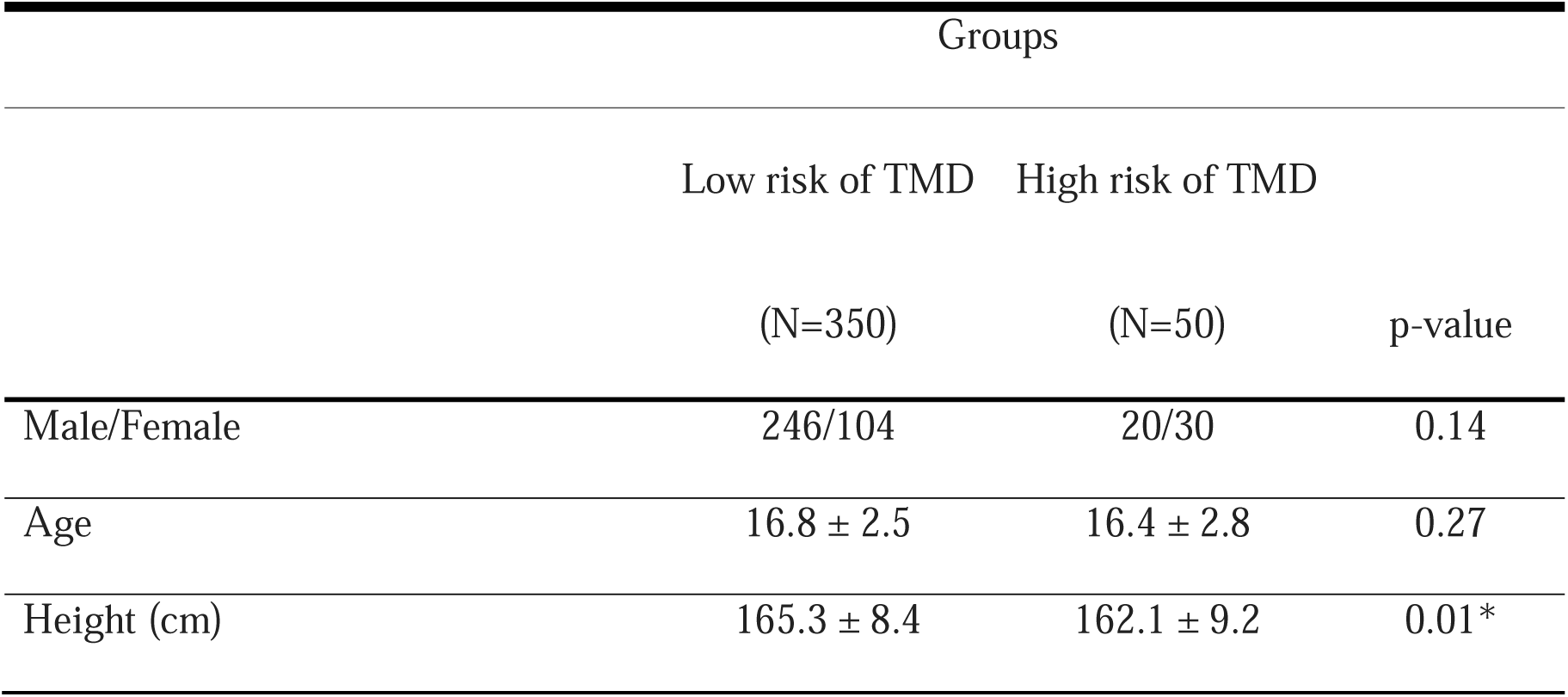
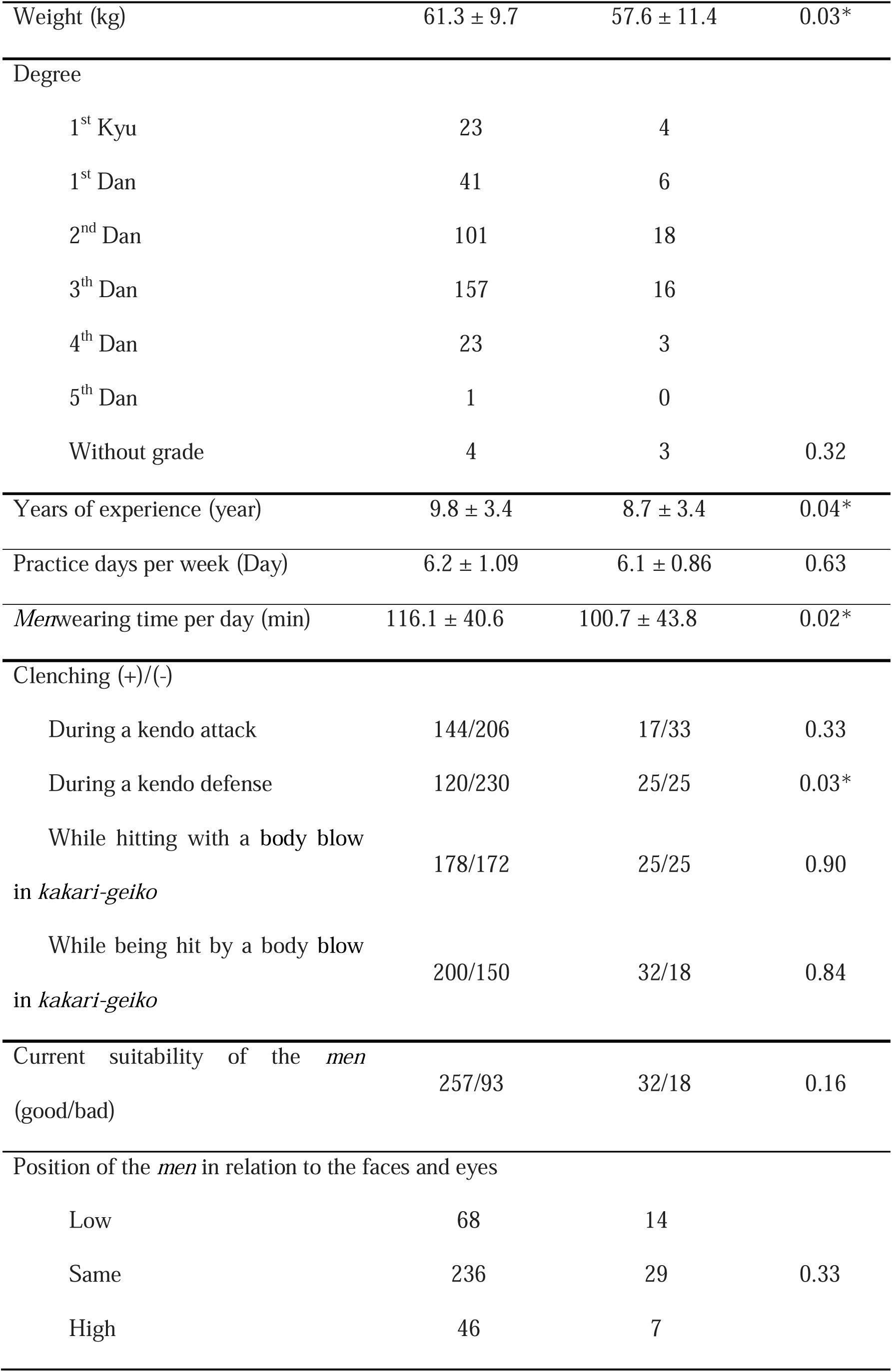
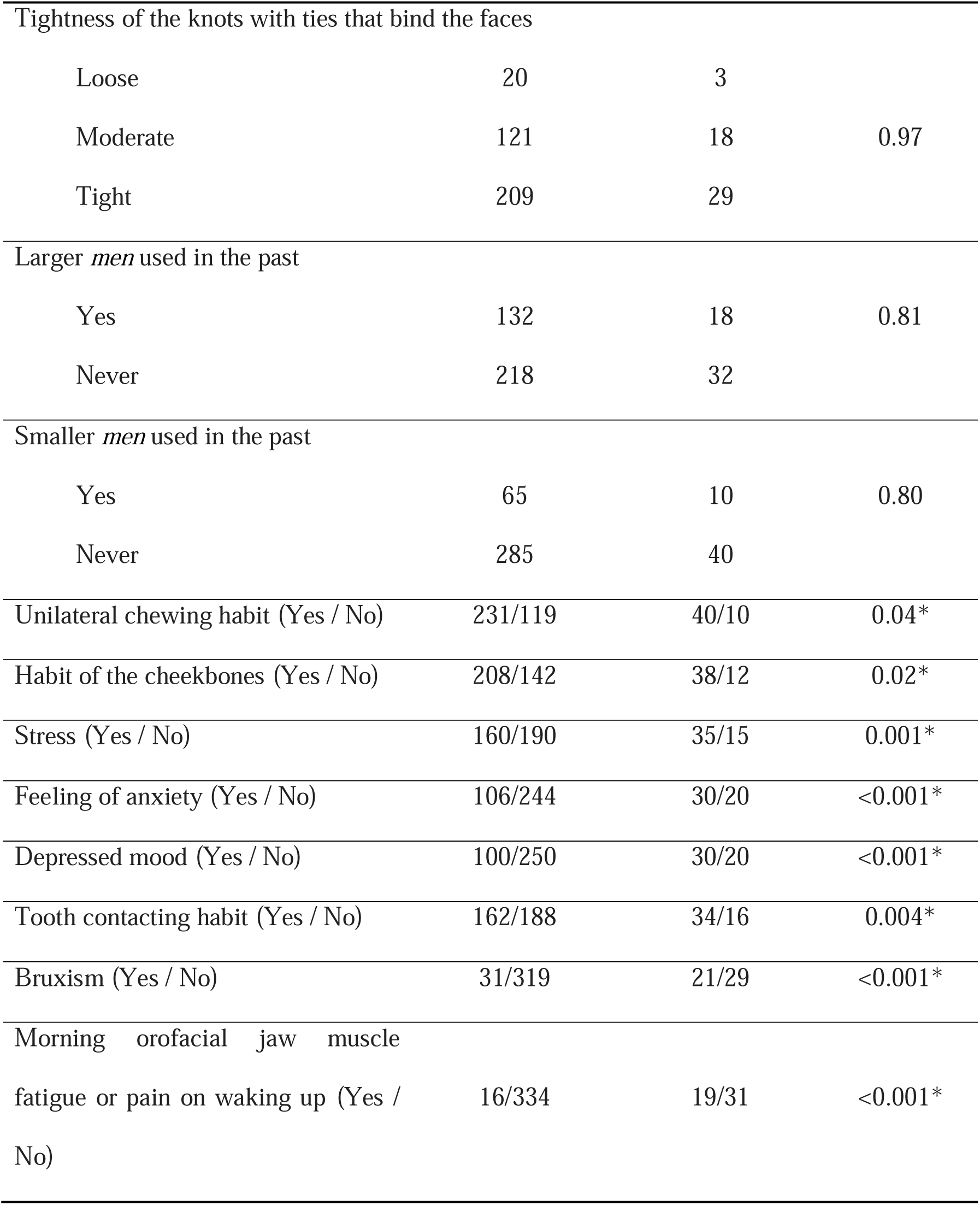
Characteristics among the high and low risk of TMD groups.

Table 6 shows the results of logistic regression analysis on oral injury and Table 7 of that on TMD. Years of experience and clenching the jaws during an offensive Kendo move significantly contributed to oral injury. Clenching the jaws during a defensive Kendo move, current *men* fit, sleep bruxism, and morning symptoms from sleep bruxism significantly contributed to TMD.

**Table 6.**
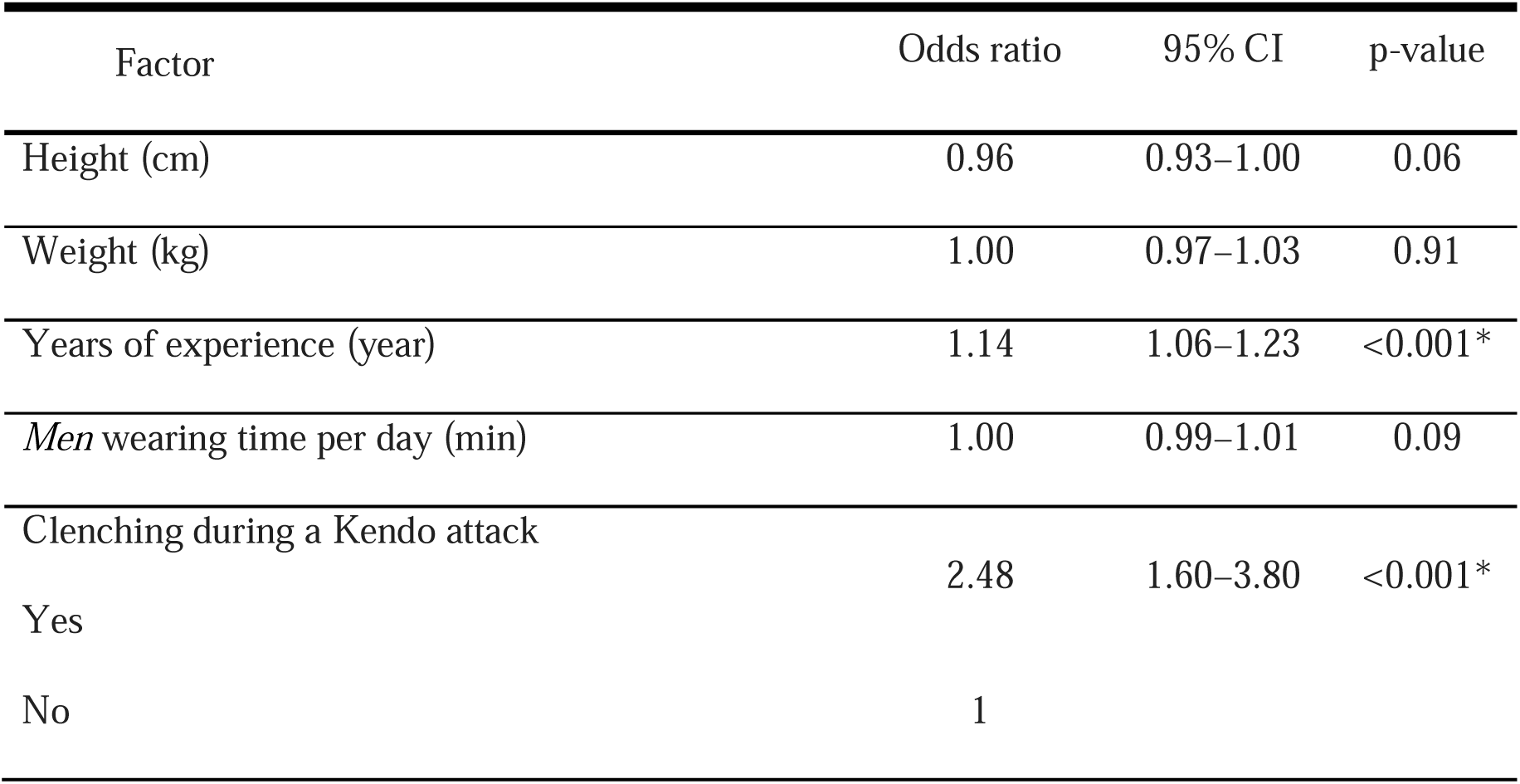
Multiple regression analysis for the trauma and non-trauma groups.

| Factor | Odds ratio | 95% CI | p-value |
| --- | --- | --- | --- |
| Height (cm) | 0.96 | 0.93–1.00 | 0.06 |
| Weight (kg) | 1.00 | 0.97–1.03 | 0.91 |
| Years of experience (year) | 1.14 | 1.06–1.23 | <0.001* |
| *Men* wearing time per day (min) | 1.00 | 0.99–1.01 | 0.09 |
| Clenching during a Kendo attack |  |  |  |
| Yes | 2.48 | 1.60–3.80 | <0.001* |
| No | 1 |  |  |

**Table 7.**
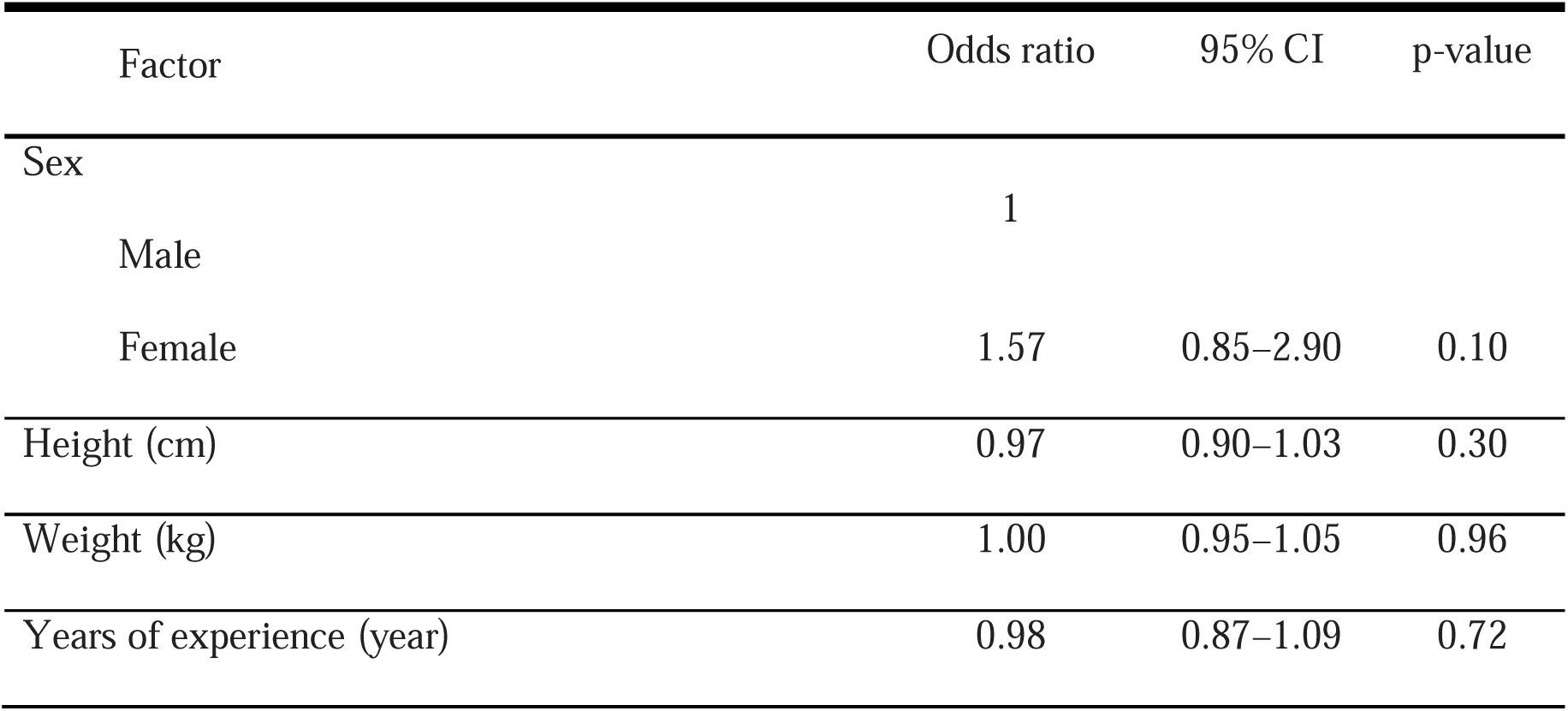
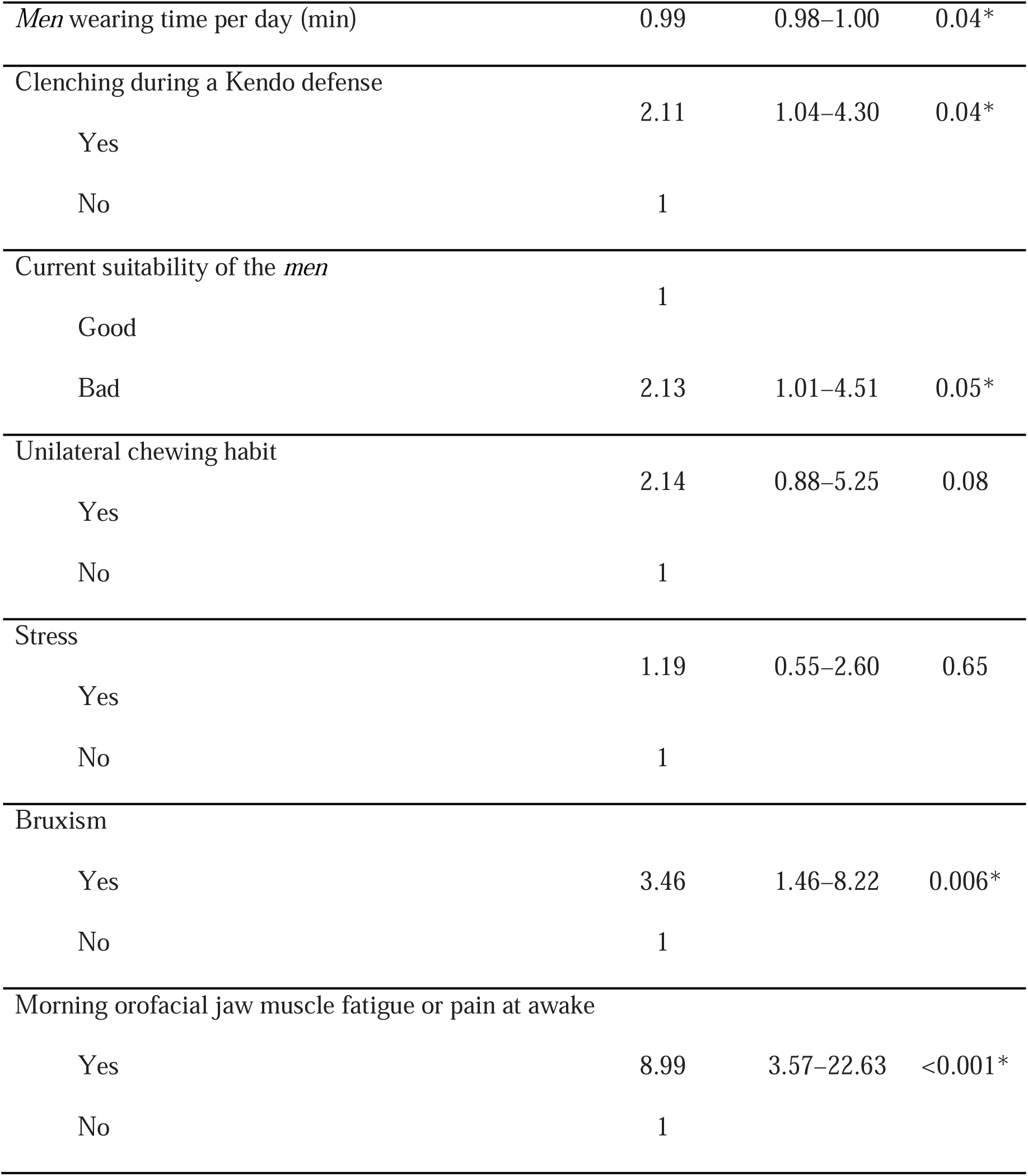
Multiple regression analysis for the high and low risk of TMD groups.

## Discussion

Previous studies have reported the injury rates associated with kendo in children. Schultzel et al. evaluated the prevalence and characteristics of injuries in Kendo [12]; 74% of men and 83% of women reported injuries to two or more body parts. The common injury sites were foot/ankle (65.1%), wrist/hand (53.5%), and elbow/forearm (48.8%). The injury types were bruising, abrasion, and strains/sprains. The injury rate during the matches was 121/1000 A–E. However, this study did not assess forehead, facial, and oral injuries associated with *men*.

Nonoyama et al. reported oral injuries in Kendo compared with other school sports [13]. The annual oral injury rate per 1000 students in Kendo for males and females was 0.62 and 0.56, respectively, among junior high school students, and 2.55 and 0.35, respectively, among high school students. This is not as high as that in other sports. However, this study only included trauma that cost more than ¥5,000 (>$40) to treat as an “injury”; thus, only relatively severe oral injuries were included in this report. Other injuries such as soft tissue trauma and TMD were not included in this study; therefore, the full extent of the oral injuries was not clear.

In the present study, other injuries, such as soft tissue trauma and TMD, were also included to obtain a detailed picture of the occurrence of oral injuries. The results showed that 44.8% of young Kendo players had a history of oral trauma. The results of logistic regression analysis with a history of oral injury as the objective variable showed that years of experience and clenching one’s jaws during a Kendo attack were significant explanatory variables (Table 6). In particular, those players who were aware of clenching during Kendo were shown to be 1.3 times more likely to have experienced an oral injury during practice. However, these data were based on an individual’s own awareness, and it is unclear whether they were actually biting down during each situation. A mouthpiece type device that detects biting and clenching in real time has been developed [14]. We believe that it is necessary to use this device to determine the actual status of biting and clenching one’s jaws during Kendo practice.

Mucosal soft tissue in the oral cavity was the most frequent site of intraoral injury. As the players always wear *men* during both competition and training in Kendo, soft tissue injury was thought to be caused by the player’s own teeth. Takeuchi et al. [15] identified that this type of injury, particularly in sports where *men* is worn, is because of the limited range of motion of the jaw resulting in accidental biting and injury to the oral mucosa. The alignment of the individual’s dentition and occlusal relationship is also thought to contribute to the occurrence of accidental biting. To clarify this relationship, it is necessary to examine the differences in the occurrence of soft tissue trauma in a prospective study classifying the participants according to their dental and occlusal relationships.

Mouth guards are rarely used in Kendo because the use of a *men* makes dental trauma less likely to occur. Nonoyama et al. [13] had shown that dental trauma in Kendo is less sever compared with that in other sports. However, mouth guards may additionally protect against soft tissue trauma. This is called the “sheathing effect” of the mouth guards, as they enclose the blade (tooth) in a sheath (mouth guard), making it safe to handle. In the past month, 32 (8.2%) Kendo players reported that their matches or training had been disrupted by dental or oral problems. Therefore, it is important to reduce the risk of accidental biting to create an environment in which players can concentrate on their competition without having to worry about soft tissue trauma.

TMD was the second most frequent intraoral injury after injury to mucosal soft tissue. The high risk of TMD group accounted for 12.5% of all participants (males, 10.9%; females, 16.1%). It has also been reported that TMD was a significant public health problem affecting approximately 5–12% of the general population [16]. According to the results of a screening questionnaire survey for TMD with 2203 participants, high risk of TMD was found in 361 (16.4%) individuals [17]. The results of our study were consistent with those of previous studies and showed that Kendo players had a similar prevalence of TMD compared to the general population.

In the present study, the results of the logistic regression analysis with TMD as the objective variable showed that clenching during a defensive Kendo move, current fit of the *men*, sleep bruxism, and morning symptoms of sleep bruxism were significant explanatory variables (Table 7). Sleep bruxism and morning symptoms of sleep bruxism are high-risk factors for TMD. In particular, the OR for TMD was shown to be 2.1 times higher among Kendo players who were aware of clenching during defense in Kendo. As the upper and lower jaws are locked in place as they clench, there is less space to buffer their impact in the maxillofacial area. Part of this may impact the TMJ. Additionally, the possibility for TMD for a player with ill-fitting *men* was 2.2 times. However, in this study, *men* fit was only subjective data, and it is not known whether it can objectively be considered to fit well or not. Therefore, it is necessary to investigate and compare the relationship with TMD after objectively evaluating the *men* fit; however, a tool that can do so objectively does not currently exist and education on how to choose the right side for oneself and the development of tools to confirm that the correct size is being used are considered necessary.

Many players who were aware of the unsuitability of the *men* took measures to improve it; the measures taken to deal with ill-fitting *men* were considered to have little impact on oral injuries such as TMD, according to the results. It is suggested that the burden on the oral injuries and TMJ could be reduced by adjusting the fit with materials such as fabric or absorbent pads.

This study found that putting on verified *men* enables proper visibility; it also reduces the risk of head fluctuating when it is hit or moving during Kendo performance, influencing the quality of performance, and thereby becoming dependent variables. Given that the length of time wearing an unverified *men* was not a potential factor associated with TMD, it is assumed that other potential factors, such as ties in the *men* and position of one’s jaws in the *men*, have worked out, regardless of the use of unverified *men*, resulting in the adjustment of the position of one’s eyes in a *men* and may possibly be an explanation.

Originally, Kendo gears had artistic influence and were made based on both esthetics and better utility, assuming that the players wore the gears correctly [18,19]. How Kendo gears affect performance is to be examined in future studies.

A limitation of this research is that the sample consisted of only students aged 18 to 22 years; ; therefore, the findings should be generalized with caution as Kendo is a sport played regardless of age, and it is highly expected that the results may vary with age. The questionnaire used in the study should be standardized and implemented to measure the differences between the youth and elderly; it is to be done via quantified methodology to fully explore the relationship between Kendo and oral trauma.

## Data Availability

The data that support the findings of this study are available from the corresponding author, [KW], upon reasonable request.

## Acknowledgments

The authors would like to thank Editage (www.editage.jp) for English language editing.

